# Concordance in the identification of pathogens by next-generation sequencing and conventional microbiology methods among neonates with culture-positive sepsis

**DOI:** 10.64898/2025.12.05.25341614

**Authors:** Divashree Jhurani, Sanjay Kumar, Varsha Mittal, Sushil Srivastava, Kirti Nirmal, Sushma Nangia, Ravinder Kaur, Pradeep Debata, Rajni Gaind, Vivek Kumar, Ramesh Agarwal, M Jeeva Sankar, Kajal Jain

## Abstract

**Introduction:** Early and accurate diagnosis of sepsis through identification of the causative pathogen is vital for the appropriate treatment of infection and avoiding misuse of antibiotics. Due to low microbial load and the presence of host DNA, earlier studies have shown poor concordance between microbiology and metagenomics results in the case of blood samples. In the present study, we standardized the protocol to obtain the concordance between pathogen(s) identified through conventional microbiological culture and those obtained through 16S metagenomic analyses for the blood and blood culture samples obtained from preterm neonates.

**Methodology:** Paired blood samples (in EDTA) and liquid blood culture samples were collected from neonates suspected of sepsis for microbial DNA extraction and conventional blood culture, respectively. Liquid blood culture samples were also used for microbial DNA extraction. DNA was extracted using Qiagen kit with additional steps of pre-extraction host DNA depletion and post extraction cleanup steps with magnetic beads. The extracted microbial DNA was further subjected to 16S rRNA sequencing using the MinION Mk1C platform.

**Results:** By using modified protocol, the concentration and quality of DNA improved. Sixteen paired samples were processed. Results obtained through metagenomics and conventional microbiology showed 100% and 87.5% concordance (at genus level) in case of liquid blood culture and blood samples respectively.

**Conclusion:** Our study compared and showed high concordance between pathogen identification in paired blood and liquid blood culture samples obtained from preterm neonates with sepsis using microbiological and molecular methods.

## Introduction

Bloodstream infections (BSIs) remain a significant cause of morbidity and mortality. Globally, it is one of the top ten causes of death (1), and among neonates, sepsis is the third leading cause of mortality (2). Quick and accurate identification of the causative pathogens is crucial for guiding appropriate treatment strategies, reducing antibiotic misuse, and improving neonatal outcomes. However, identifying pathogens from conventional blood cultures takes >72 hours in most settings. Also, it is challenging to obtain enough volume of blood from a sick neonate specially when born preterm. This leads to sterile cultures and further delays the treatment. To overcome the challenges faced by the conventional blood culture method, timely and accurate identification of the causative pathogens using next-generation sequencing (NGS) techniques has been explored (3).

The advent of NGS technologies has revolutionized the field of metagenomics, enabling researchers to unravel the diverse microbial communities inhabiting various environments. NGS works well with sample types like feces, swabs, lavage, pus and CSF (4–7). However, clinical diagnosis of sepsis depends on the blood culture. Due to its very low microbial load and the presence of host DNA, blood is not considered an appropriate sample for pathogen identification using NGS techniques. Studies have shown very poor concordance between conventional culture and NGS results in the case of blood samples (8–10). Concordance ranged from 0 to 25% among patients with culture-positive sepsis in these studies. One recent study showed no concordance in the identification of bacteria by metagenomic shotgun sequencing and blood cultures and hence suggested further optimization of sample preparation methods (11).

Rapid and accurate pathogen identification is crucial for timely treatment in case of sepsis. NGS offers promise in this area, although challenges persist in processing blood samples due to low microbial load and host DNA interference (12). In the current study, we employed a few modifications in the blood sample processing to improve the quality of microbial DNA extracted and accurately identify the pathogen from the blood sample using NGS to achieve concordance with the conventional microbiology results.

## Methodology

### Sample collection and processing

Anonymized samples from an ongoing multi-centric study on neonatal sepsis were prospectively collected from three clinical sites and transported to the nodal centre. At each clinical site, on clinical suspicion, the dedicated research personnel collected blood from the preterm neonates (28 to 34 weeks’ gestation) before initiation of antibiotics using aseptic procedures after obtaining consent from the parents. 1 mL of blood sample was put in BACTEC culture bottles for microbiology culture, while another 0.5 mL of blood sample was collected in an EDTA vial (B1). The blood culture bottles with blood samples were placed in an automated culture system and monitored for ‘beep’. From these incubated culture bottles, 0.5 mL sample was collected within 10 hours window. This sample mixture collected from the incubated culture bottle was labelled as ‘liquid culture’ sample (B2). Hence, paired samples (B1 and B2) were obtained from each neonate for microbial DNA extraction and NGS: one blood sample in EDTA (B1) and another after incubation in the culture bottles (B2) (figure 1).

**Figure 1:**
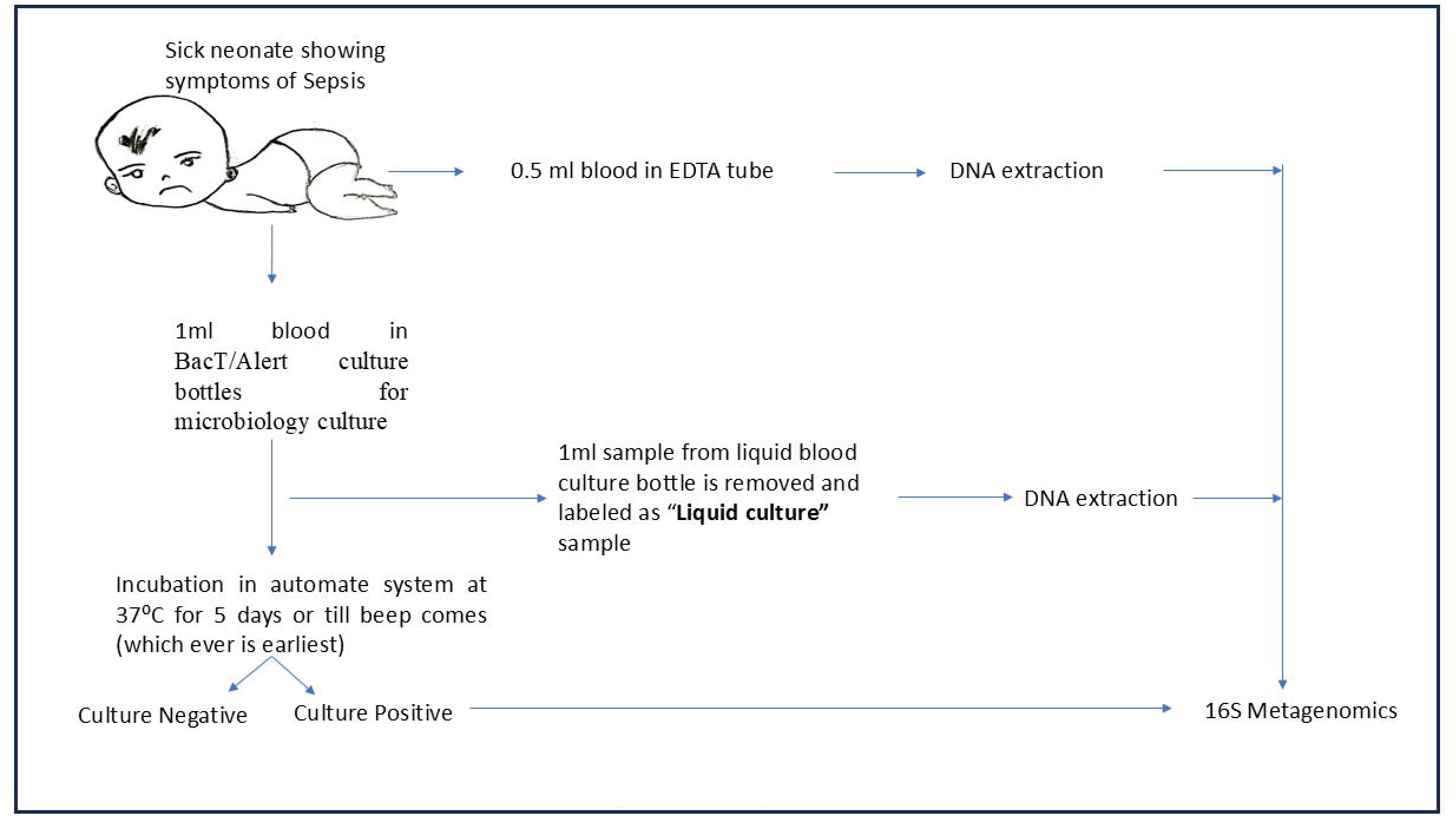
Sample flow

The incubated bottles on ‘beep’ were cultured on 5% sheep blood agar (Biomeriux) and MacConkey agar (Biomeriux or Hi-media) and incubated at 37°C for 18 to 24 hours. Routine microbiology methods were used to identify the pathogen. This was followed by confirmation of identification of the pathogen using MALDI-TOF.

### DNA extraction from blood samples

Processing of samples was done within 15 days after collection; until that samples were stored at −80 °C. Samples that were confirmed to be culture positive by microbiology methods were processed further for DNA extraction. For DNA extraction 400 µl of the sample was mixed with 20% saponin, and then incubated at 37°° C for 30 minutes. After this, mechanical disruption of the bacterial cell wall was done by transferring the sample into the Pathogen lysis tube (S) and bead beating for 2 minutes at 50 Hz using the Qiagen Tissue Lyser LT. The supernatant from the tube was transferred to the new tube and incubated with 15 µl of Metapolyzyme (10 µg/µl) at 37 °C for an hour. After these pre-processing steps, extraction of DNA was done using the Qiagen DNeasy Blood and Tissue DNA Kit (69504) as per the manufacturer’s instructions. DNA QC were determined using the Qubit fluorometric method and nanodrop. For DNA extracted from liquid blood culture, additional step for cleaning the DNA with AMPure XP (Beckman Coulter) beads gave better quality of DNA.

Clean-up of DNA using beads was performed as per manufacturer’s protocol. (Supplementary figure 1)

### 16S rRNA sequencing using ONT Mk1C

For 16S rRNA sequencing, the first step was to amplify the V1 to V9 segment of 16S rRNA; this was done using conventional PCR for 16S rRNA using the extracted DNA as a template. For this purpose, 25 µl of reaction was prepared using 12.5 µl of 2x Go Taq master mix, 2 µl of forward and reverse primers (5 pmol each), and 50–70 ng of DNA template. Amplification was performed on a Veriti 96-well thermal cycler by Applied Biosciences using the following PCR conditions: 94 °C x 2 mins, then 35 cycles of 94 °C x 45 sec, 53 °C x 45 sec, and 72 °C x 2 mins and a final extension step of 72 °C x 10 mins. Primers used in this PCR reaction were universal 27F (5′-AGAGTTTGATCCTGGCTCAG-3′) and 1492R (5′-TACGGYTACCTTGTTACGACTT-3′). For 2^nd^ PCR, 1 µl of conventional PCR product was used as a template for amplification using SQK-16S024 kit (with barcoded primers).

### Library Preparation

The PCR products of the second PCR underwent purification using AMPure XP beads (Beckman) and were eluted in 10 µl of elution buffer. The final elute of 10 µl was taken for library preparation for 16S metagenomics and every step of library preparation, including loading and sequencing, was conducted in accordance with the 16S sequencing protocol provided by Oxford Nanopore Technologies (ONT). The data obtained by sequencing was analysed using the online software EPI2ME (Metrichor™ support), a cloud-based data-analysis platform developed by Metrichor Ltd for Oxford Nanopore Technologies.

## Results

Among 17 preterm neonates with positive blood cultures, paired samples (B1 and B2) were obtained from 16 neonates. In one neonate, only the liquid culture sample (B2) could be obtained. Subsequently, 16S rRNA NGS was conducted for all 33 samples. Microbiology cultures revealed *Klebsiella pneumoniae* in 10 cases, coagulase-negative staphylococci in 4 cases, *Acinetobacter baumannii* in 2 and *Proteus mirabilis* in 1.

Details regarding quality control checks of extracted DNA from both blood and liquid culture samples have been provided in supplementary table 1. DNA concentration from blood and liquid blood culture samples ranged from 18-312 ng/µl and 24-161 ng/µl, respectively. The 260/280 ratio for blood samples was between 1.2 and 1.8; and for liquid blood culture samples, between 0.2 and 0.6. After the additional clean-up protocol for liquid blood culture samples, a significant enhancement in DNA quality was observed, and the 260/280 ratio increased to 1-1.6.

The concordance of 16S NGS with culture results was considered if the same organism (at genus level) isolated from culture was among the five most abundant organisms in a sample. EPI2ME results from the blood sample showed 87.5% concordance (14/16). However, the number of reads per sample was less in most cases. Liquid culture samples showed 100% concordance with the microbiology results at the genus level (Table 1) and 16/17 (94.1%) concordance at the species level (Supplementary table 2). The EPI2ME output at genus sample from two neonates (DNA extracted from blood sample and liquid culture sample) are depicted in Supplementary figure 2.

**Table 1:** Concordance to identify pathogenic bacteria (at genus level) between 16S rRNA NGS and organism grown in microbiological culture for each sample.

| Sample ID | Microbiology culture results | 16S results from liquid culture samples (number of reads) | 16S results from blood samples (number of reads) |
| --- | --- | --- | --- |
| 11397 | Mixed CoNS | *Staphylococcus* (1,674)<br>*Bacillus* (5)<br>*Macrococcus* (4)<br>*Serratia* (2)<br>*Elizabethkingia* (1) | *Pseudomonas* (14)<br>*Alteromonas* (11)<br>*Serratia* (9)<br>*Acinetobacter* (7)<br>*Enterobacter* (3)<br>*Staphylococcus* (2) |
| 11402 | *Acinetobacter baumannii* | **Acinetobacter (3,351)**<br>*Stutzerimonas* (9)<br>*Staphylococcus* (7)<br>*Serratia* (6)<br>*Pseudomonas* (6) | *Sample Not Received* |
| 20790 | *Proteus mirabilis* | **Proteus (9,006)**<br>*Xenorhabdus* (333)<br>*Acinetobacter* (53)<br>*Yersinia* (20)<br>*Enterobacter* (16) | **Proteus (2,305)**<br>*Xenorhabdus* (37)<br>*Sodalis* (5)<br>*Oceanisphaera* (3)<br>*Edwardsiella* (3) |
| 20806 | *Acinetobacter baumannii* | **Acinetobacter (3)**<br>*Klebsiella* (2) | *Proteus* (42)<br>**Acinetobacter (37)**<br>*Streptococcus* (13)<br>*Pseudomonas* (9)<br>*Enterobacter* (7) |
| 20810 | *Staphylococcus haemolyticus* | **Staphylococcus (9)** | *Acinetobacter* (19)<br>*Pseudomonas* (10)<br>*Proteus* (7)<br>*Paracoccus* (6)<br>**Staphylococcus (6)** |
| 20819 | *Klebsiella pneumoniae* | **Klebsiella (1,994)**<br>*Enterobacter* (37)<br>*Kosakonia* (12)<br>*Citrobacter* (4)<br>*Xenorhabdus* (4) | *Acinetobacter* (19)<br>**Klebsiella (14)**<br>*Pseudomonas* (9)<br>*Enterobacter* (7)<br>*Staphylococcus* (6) |
| 40307 | *Staphylococcus hominis* | **Staphylococcus (1,103)**<br>*Pseudomonas* (14)<br>*Serratia* (8)<br>*Bacillus* (6)<br>*Listeria* (3) | *Acinetobacter* (5)<br>*Klebsiella* (4)<br>**Staphylococcus (4)**<br>*Christiangramia* (1)<br>*Chryseobacterium* (1) |
| 40309 | *Klebsiella pneumoniae* | **Klebsiella (1,612)**<br>*Enterobacter* (46)<br>*Kosakonia* (15)<br>*Pectobacterium* (7)<br>*Staphylococcus* (3) | **Klebsiella (70)**<br>*Enterobacter* (28)<br>*Pseudomonas* (12)<br>*Tatumella* (8)<br>*Acinetobacter* (8) |
| 40323 | *Staphylococcus epidermidis* | **Staphylococcus (1,264)**<br>*Bacillus* (7)<br>*Serratia* (3)<br>*Pseudomonas* (3)<br>*Macroccoccus* (3) | *Pseudomonas* (35)<br>*Acinetobacter* (14)<br>*Klebsiella* (7)<br>*Streptococcus* (7)<br>*Serratia* (5)<br>**Staphylococcus (4)** |
| 40325 | *Klebsiella pneumoniae* | **Klebsiella (1,143)**<br>*Enterobacter* (47)<br>*Enterococcus* (42)<br>*Kosakonia* (11)<br>*Pectobacterium* (5) | *Pseudomonas* (20)<br>**Klebsiella (18)**<br>*Acinetobacter* (11)<br>*Streptococcus* (8)<br>*Enterobacter* (4) |
| 40315 | *Klebsiella pneumoniae* | **Klebsiella (1,507)**<br>*Enterobacter* (37)<br>*Kosakonia* (14)<br>*Pectobacterium* (10)<br>*Cedecea* (2) | **Klebsiella (507)**<br>*Enterobacter* (45)<br>*Pseudomonas* (29)<br>*Serratia* (10)<br>*Acinetobacter* (16) |
| 40333 | *Klebsiella pneumoniae* | *Enterococcus* (1,528)<br>***Klebsiella*** (177)<br>*Staphylococcus* (120)<br>*Enterobacter* (7)<br>*Carnobacterium* (5) | *Acinetobacter* (21)<br>*Pseudomonas* (13)<br>*Staphylococcus* (8)<br>***Klebsiella*** (5)<br>*Streptococcus* (3) |
| 40372 | *Klebsiella pneumoniae* | ***Klebsiella*** (44,127)<br>*Enterobacter* (1,575)<br>*Kosakonia* (355)<br>*Pectobacterium* (145)<br>*Tatumella* (60) | ***Klebsiella*** (1,249)<br>*Bacillus* (75)<br>*Enterobacter* (25)<br>*Kosakonia* (6)<br>*Pectobacterium* (3) |
| 40377 | *Klebsiella pneumoniae* | ***Klebsiella*** (1,733)<br>*Enterobacter* (40)<br>*Kosakonia* (14)<br>*Citrobacter* (2)<br>*Pectobacterium* (2) | ***Klebsiella*** (32)<br>*Acinetobacter* (13)<br>*Streptococcus* (8)<br>*Enterobacter* (6)<br>*Proteus* (6) |
| 40378 | *Klebsiella pneumoniae* | ***Klebsiella*** (1,246)<br>*Enterobacter* (40)<br>*Kosakonia* (15)<br>*Pectobacterium* (5)<br>*Tatumella* (3) | *Serratia* (7)<br>*Acinetobacter* (6)<br>*Pseudomonas* (5)<br>***Klebsiella*** (4)<br>*Staphylococcus* (3) |
| 40379 | *Klebsiella pneumoniae* | ***Klebsiella*** (1,556)<br>*Enterobacter* (55)<br>*Kosakonia* (10)<br>*Pectobacterium* (8)<br>*Pseudenterobacter* (2) | *Pseudomonas* (15)<br>*Staphylococcus* (12)<br>***Klebsiella*** (11)<br>*Acinetobacter* (10)<br>*Enterobacter* (6) |
| 40376 | *Klebsiella pneumoniae* | ***Klebsiella*** (2,416)<br>*Enterobacter* (108)<br>*Kosakonia* (18)<br>*Citrobacter* (5)<br>*Pectobacterium* (5) | *Pseudomonas* (32)<br>*Serratia* (16)<br>***Klebsiella*** (8)<br>*Staphylococcus* (7)<br>*Acinetobacter* (6) |

## Discussion

Earlier studies have shown less concordance of 16S rRNA NGS with microbiology methods, specifically from blood samples (8–10). Remarkable concordance was observed between the two methods in our study, indicating that the modifications made to the blood sample processing protocol have worked well and efficiently identified the pathogen, as found through microbiological culture.

We processed paired samples of both blood specimens and liquid blood cultures from culture-positive neonates. Our protocol included timely processing of blood samples, host DNA removal through saponin, mechanical (bead beating) and enzymatic lysis, and incorporation of further purification steps post-extraction to enhance DNA quality, particularly for liquid blood culture samples. It was also seen that increasing the duration/ intensity of the mechanical lysis step fragmented the DNA, making it unavailable for the whole-length 16S PCR. Nested PCR was employed to increase the concentration of the target amplicon, thus obtaining the post-PCR Qubit concentrations as required for the ONT sequencing experiment. This improved DNA quality which likely contributed to the higher accuracy of NGS results.

Plasma has been used in earlier studies for pathogen detection. We preferred blood over plasma as the sensitivity of 16S rRNA NGS from plasma is limited due to the low amount of microbial DNA in plasma and the short half-life of cfDNA (13). Blood has less bacterial load, so incubating the blood sample in a culture bottle for a few hours helped in the enrichment of the sample. Interestingly, this gave a substantially high number of pathogen reads in these samples compared to the blood samples, where the number of reads of the pathogen was similar to the background contamination, making it challenging to identify the true pathogen. A recently published study also showed high accuracy (100%) for bacterial and fungal microorganisms using the same strategy of blood culture fluid for the mNGS testing (14). However, in our study, we collected the sample for NGS within 10 hours, instead of waiting for the beep, which may help in earlier identification. We chose only culture-positive samples to understand the concordance for identification of pathogen. Hence it delayed the metagenomic process as it required confirmation of microbiology results. But once validated, this process holds promise for early identification of pathogens, with no extra sample volume required.

A long-read platform (MinION Mk1C) was used for sequencing. Long-read sequencing helps in increased taxonomic classification and accuracy due to its ability to read the long fragments and cover the entire region of the 16S rRNA (15). The strength of our study is the rigorous methodology followed and the complete concordance with the microbiology results. The limitation of our study is the smaller sample size and absence of culture-negative samples.

However, the observations from the study have raised some pertinent questions. As seen, that in few blood samples there were more than one bacteria (that are known to be pathogenic), but after incubation, only one bacteria is getting multiplied. Does it refer to polymicrobial sepsis or perhaps culture has low sensitivity which leads to poor choice of antibiotics. This poorly informed choice of antibiotics may drive the emergence and/or selection of MDR and XDR pathogens that lead to higher morbidity and mortality.

## Conclusion

In conclusion, this study highlights the potential of NGS techniques, when combined with optimized blood sample processing, to revolutionize the diagnosis of sepsis. This will help in early identification of pathogens in sepsis. Further research and validation studies are required to assess the applicability of this approach in a broader clinical context.

## Data Availability

All data produced in the present work are contained in the manuscript

## Abbreviations

16S rRNA: 16S ribosomal ribonucleic acid
BSI: Blood stream infections
CSF: Cerebrospinal fluid
cfDNA: Cell Free Deoxyribonucleic Acid
EDTA: Ethylenediaminetetraacetic acid
DNA: Deoxyribonucleic Acid
ONT: Oxford Nanopore Technologies
MDR: Multi Drug Resistant
mNGS: Metagenomic Next Generation Sequencing
NGS: Next Generation Sequencing
MALDI-TOF: Matrix Assisted Laser Desorption Ionization-time of flight
PCR: Polymerase Chain Reaction
XDR: Extensively Drug Resistant

## Conflict of interest

The authors have no conflict of interest relevant to this article to disclose.

## Funding information

This work was funded by the Department of Biotechnology, Government of India *(BT/PR38173/MED/97/474/2020)*.

## Ethics approval

The Institutional Ethics Committee of AIIMS and three participating clinical sites (*Ethics approval ref no. IEC-1087/06*.*11*.*2020, RP-42/2020*); and Institutional Committee on Biosafety (*IBSC approval ref no. IBSC 0521_KJ)* approved the study. The work described has been carried out in accordance with The Code of Ethics of the World Medical Association (Declaration of Helsinki).

## Availability of data and material

Data will be available on reasonable request.

## Acknowledgement

We acknowledge the families who gave consent to participate in the study. We also acknowledge the teams involved at the three clinical sites.

## Author Contributions

KJ, DJ, VK, MJS, RA: designed the study and prepared the manuscript

KJ, DJ, SK: implemented the study

SS, SN, PD: enrolled neonates at the clinical sites

KN, RK, RG: supervised conventional microbiology at clinical sites

KJ, DJ, MJS: supervised the implementation of the study and finalized the manuscript.

DJ, SK: Performed the wet lab experiments of the study

DJ, VM, KJ: Did the analysis

## Notes

### Competing Interest Statement

The authors have declared no competing interest.

### Funding Statement

This study was funded by Department of Biotechnology grant number BT/PR38173/MED/97/474/2020

### Author Declarations

The Institutional Ethics Committee of AIIMS and three participating clinical sites (Ethics approval ref no. IEC-1087/06.11.2020, RP-42/2020); and Institutional Committee on Biosafety (IBSC approval ref no. IBSC 0521_KJ) approved the study.

