## Supplementary data for "Concordance in the identification of pathogens by next-generation sequencing and conventional microbiology methods among neonates with culture-positive sepsis"

**Methodology Details**

**Supplementary table 1**: Quality check of extracted DNA from samples

|  |  | **Post-extraction and column washing** | | **Post-extraction and column washing and further clean-up with beads** | |
| --- | --- | --- | --- | --- | --- |
| **Sample ID** | **Sample Type** | **Concentration**  **(ng/µl)** | **260/280** | **Concentration**  **(ng/µl)** | **260/280** |
| 11397 | Liquid culture | **161** | **0.5** | **228** | **1** |
|  | Blood | 72 | 1.5 |  |  |
| 11402 | Liquid culture | **76** | **0.3** | **68** | **1.1** |
| 20790 | Liquid culture | **87** | **0.3** | **210** | **1.5** |
|  | Blood | 33 | 1.5 |  |  |
| 20806 | Liquid culture | **86** | **0.2** | **92** | **1** |
|  | Blood | 20 | 1.2 |  |  |
| 20810 | Liquid culture | **89** | **0.2** | **71** | **1** |
|  | Blood | 30 | 1.5 |  |  |
| 20819 | Liquid culture | **67** | **0.2** | **87** | **1** |
|  | Blood | 18 | 1.4 |  |  |
| 40307 | Liquid culture | **46** | **0.3** | **15** | **0.7** |
|  | Blood | 63 | 1.6 |  |  |
| 40309 | Liquid culture | **56** | **0.6** | **75** | **1.5** |
|  | Blood | 61 | 1.5 |  |  |
| 40323 | Liquid culture | **75** | **0.3** | **53** | **0.9** |
|  | Blood | 312 | 1.7 |  |  |
| 40325 | Liquid culture | **32** | **0.5** | **52** | **1.5** |
|  | Blood | 174 | 1.7 |  |  |
| 40315 | Liquid culture | **37** | **0.4** | **54** | **1.3** |
|  | Blood | 172 | 1.7 |  |  |
| 40333 | Liquid culture | **31** | **1** | **95** | **1.6** |
|  | Blood | 70 | 1.3 |  |  |
| 40372 | Liquid culture | **29** | **1** | **92** | **1.3** |
|  | Blood | 46 | 1.6 |  |  |
| 40377 | Liquid culture | **53** | **0.3** | **138** | **1.3** |
|  | Blood | 24 | 1.7 |  |  |
| 40378 | Liquid culture | **29** | **0.5** | **55** | **1.6** |
|  | Blood | 91 | 1.8 |  |  |
| 40379 | Liquid culture | **130** | **0.3** | **141** | **1.3** |
|  | Blood | 75 | 1.2 |  |  |
| 40376 | Liquid culture | **110** | **0.2** | **171** | **1.4** |
|  | Blood | 223 | 1.7 |  |  |

**Supplementary table 2**: Concordance to identify pathogenic bacteria (at species level) between 16S rRNA NGS and organism grown in microbiological culture

| **Sample ID** | **Microbiology culture results** | **16S results from liquid culture samples (number of reads)** | **16S results from blood samples (number of reads)** |
| --- | --- | --- | --- |
| 20790 | *Proteus mirabilis* | ***Proteus mirabilis (8,880)***  *Xenorhabdus hominickii (229)*  *Proteus terrae (42)*  *Proteus vulgaris (40)*  *Proteus hauseri (23)* | ***Proteus mirabilis (2,259)***  *Proteus terrae (25)*  *Xenorhabdus hominickii (19)*  *Proteus vulgaris (11)*  *Proteus hauseri (8)* |
| 20806 | *Acinetobacter baumannii* | ***Acinetobacter baumannii (3)***  *Klebsiella pneumoniae (2)* | *Proteus mirabilis (42)*  ***Acinetobacter baumannii (6)*** *Cutibacterium acnes (6)*  *Klebsiella pneumoniae (5) Acinetobacter kanungonis (5)* |
| 20810 | *Staphylococcus haemolyticus* | *Staphylococcus taiwanensis (7)* *Staphylococcus borealis (1)*  *Staphylococcus saprophyticus (1)* | *Proteus mirabilis (7)*  *Acinetobacter soli (5)*  *Klebsiella pneumoniae (4) Acinetobacter vivianii (4) Cutibacterium acnes (4)* |
| 20819 | *Klebsiella pneumoniae* | ***Klebsiella pneumoniae (1,932)***  *Klebsiella quasivariicola (43)*  *Enterobacter cloacae (32)*  *Klebsiella aerogenes (15)*  *Kosakonia radicincitans (9)* | ***Klebsiella pneumoniae (8)*** *Pseudomonas juntendi (4) Enterobacter sichuanensis (3) Klebsiella aerogenes (3)*  *Klebsiella quasipneumoniae (3)* |
| 40307 | *Staphylococcus hominis* | ***Staphylococcus hominis (993)*** *Staphylococcus ratti (24)*  *Staphylococcus taiwanensis (19) Staphylococcus borealis (18)*  *Serratia liquefaciens (6)* | *Klebsiella pneumoniae (4) Staphylococcus ratti (2) Staphylococcus roterodami (2) Christiangramia antarctica (1) Chryseobacterium ureilyticum (1)* |
| 40309 | *Klebsiella pneumoniae* | ***Klebsiella pneumoniae (1,438)***  *Klebsiella quasipneumoniae (118)*  *Enterobacter cloacae (39)*  *Klebsiella aerogenes (33)*  *Kosakonia sacchari (12)* | ***Klebsiella pneumoniae (52)*** *Enterobacter quasiroggenkampii (17) Tatumella terrea (8)*  *Enterobacter cloacae (5)*  *Klebsiella aerogenes (5)* |
| 40323 | *Staphylococcus epidermidis* | *Staphylococcus saccharolyticus (362)*  *Staphylococcus arlettae (352)* ***Staphylococcus epidermidis (216)***  *Staphylococcus hominis (176) Staphylococcus capitis (30)* | *Pseudomonas fluorescens (10) Pseudomonas oligotrophica (6) Klebsiella pneumoniae (5) Rhodocyclus gracilis (4)*  *Acinetobacter indicus (4)* |
| 40325 | *Klebsiella pneumoniae* | ***Klebsiella pneumoniae (1,028 )***  *Klebsiella quasipneumoniae (69) Enterococcus faecium (37)*  *Klebsiella aerogenes (30)*  *Enterobacter cloacae (29)* | ***Klebsiella pneumoniae (10)***  *Pseudomonas oligotrophica (6) Pseudomonas protegens (4) Rhodocyclus gracilis (3)*  *Sodalis ligni (3)* |
| 40315 | *Klebsiella pneumoniae* | ***Klebsiella pneumoniae (1,357)***  *Klebsiella quasipneumoniae (98)*  *Klebsiella aerogenes (30)*  *Enterobacter cloacae (24)*  *Kosakonia sacchari (13)* | ***Klebsiella pneumoniae (376)***  *Klebsiella aerogenes (66)*  *Klebsiella quasipneumoniae (34) Enterobacter cloacae (31)*  *Klebsiella pasteurii (18)* |
| 40333 | *Klebsiella pneumoniae* | *Enterococcus faecalis (1,471)*  ***Klebsiella pneumoniae (166)***  *Staphylococcus taiwanensis (79) Staphylococcus borealis (14)*  *Staphylococcus haemolyticus (10)* | *Acinetobacter septicus (5) Acinetobacter soli (4)*  ***Klebsiella pneumoniae (3)*** *Acinetobacter refrigeratoris (2) Pseudomonas juntendi (2)* |
| 40372 | *Klebsiella pneumoniae* | ***Klebsiella pneumoniae (37,086)***  *Klebsiella quasipneumoniae (4,217)*  *Klebsiella aerogenes (1,658)*  *Enterobacter cloacae (1,233)*  *Klebsiella pasteurii (687)* | ***Klebsiella pneumoniae (1,118)***  *Klebsiella quasipneumoniae (94)*  *Bacillus tropicus (64)*  *Klebsiella aerogenes (24)*  *Enterobacter cloacae (14)* |
| 40377 | *Klebsiella pneumoniae* | ***Klebsiella pneumoniae (1,532)***  *Klebsiella quasipneumoniae (137)*  *Klebsiella aerogenes (48)*  *Enterobacter cloacae (33)*  *Kosakonia sacchari (10)* | ***Klebsiella pneumoniae (18)***  *Klebsiella aerogenes (7)*  *Proteus mirabilis (5)*  *Klebsiella quasipneumoniae (4) Acinetobacter soli (4)* |
| 40378 | *Klebsiella pneumoniae* | ***Klebsiella pneumoniae (1,122)***  *Klebsiella quasipneumoniae (90) Enterobacter cloacae (29)*  *Klebsiella aerogenes (24)*  *Kosakonia sacchari (10)* | *Serratia liquefaciens (4)*  *Serratia quinivorans (3)*  *Weissella cibaria (3)*  ***Klebsiella pneumoniae (2)*** *Acinetobacter kanungonis (2)* |
| 40379 | *Klebsiella pneumoniae* | ***Klebsiella pneumoniae (1,273)***  *Klebsiella quasipneumoniae (171)*  *Klebsiella aerogenes (68)*  *Enterobacter cloacae (43)*  *Klebsiella pasteurii (19)* | ***Klebsiella pneumoniae (6)***  *Klebsiella aerogenes (4)*  *Pseudomonas protegens (4) Stutzerimonas stutzeri (4) Enterobacter sichuanensis (3)* |
| 40376 | *Klebsiella pneumoniae* | ***Klebsiella pneumoniae (1,991)***  *Klebsiella quasipneumoniae (264) Enterobacter cloacae (89)*  *Klebsiella aerogenes (88)*  *Klebsiella pasteurii (44)* | *Serratia liquefaciens (13)*  ***Klebsiella pneumoniae (7)*** *Pseudomonas protegens (7) Pseudomonas fluorescens (6) Staphylococcus ratti (6)* |
| 11397 | *Mixed CONS* | *Staphylococcus taiwanensis (1,270) Staphylococcus borealis (156) Staphylococcus haemolyticus (109) Staphylococcus ratti (43)*  *Staphylococcus devriesei (11)* | *Alteromonas mediterranea (11) Serratia liquefaciens (6)*  *Pseudomonas oligotrophica (4) Serratia quinivorans (3)*  *Pseudomonas chlororaphis (2)* |
| 11402 | *Acinetobacter baumannii* | ***Acinetobacter baumannii (3,170)*** *Acinetobacter kanungonis (29) Acinetobacter oleivorans (23)*  *Acinetobacter vivianii (17)*  *Acinetobacter sichuanensis (15)* | *Sample not received* |

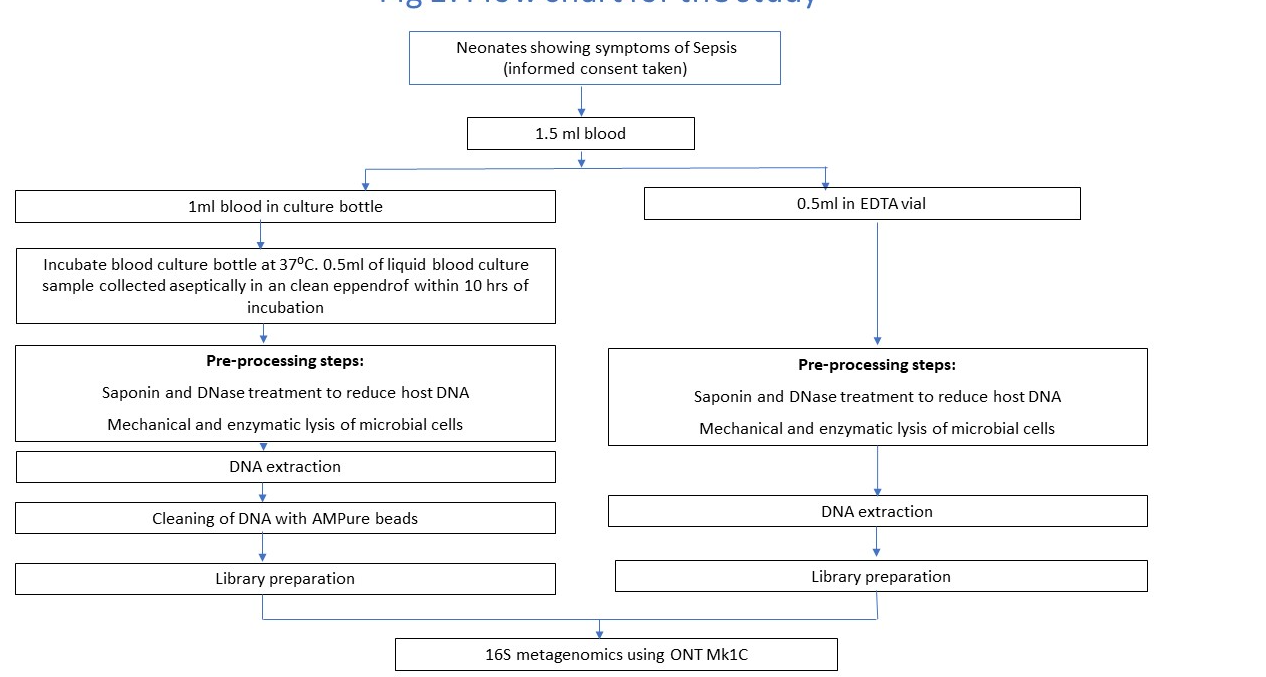

**Supplementary figure 1: Sample processing steps**

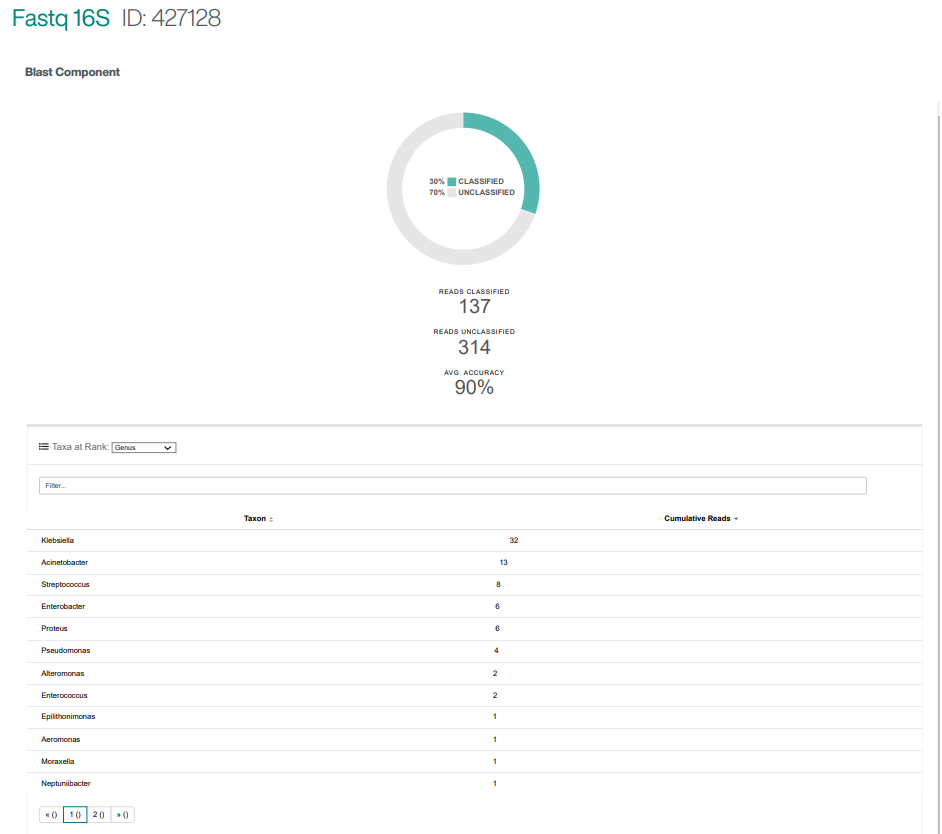

**A**

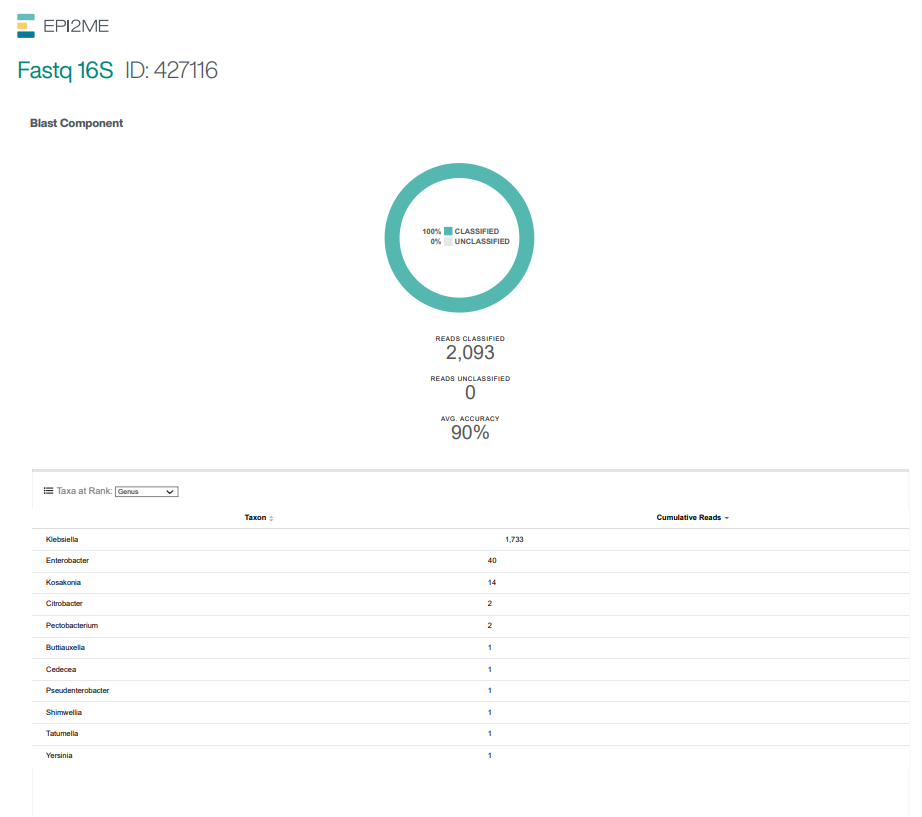

**B**

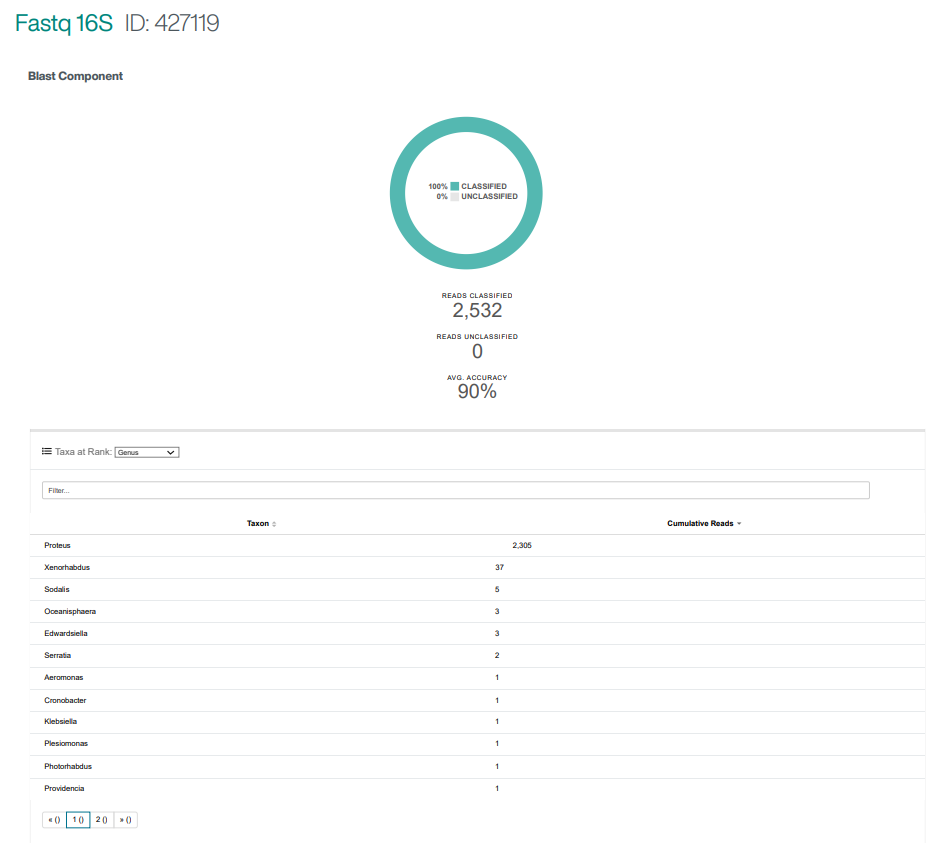

**C**

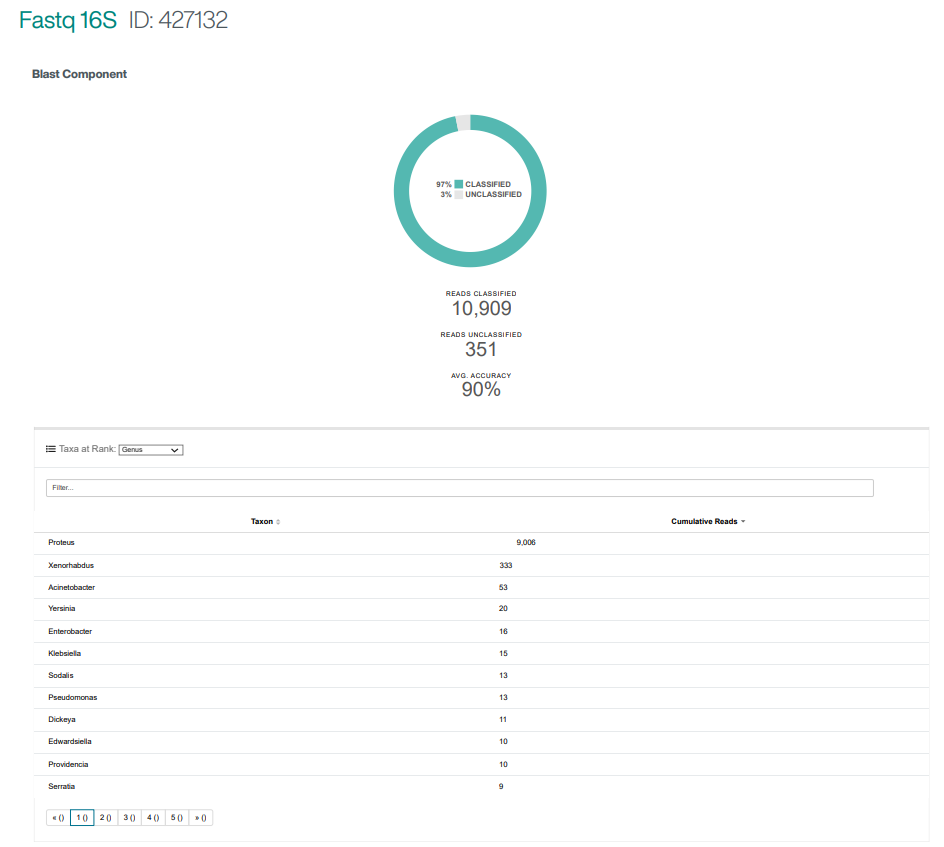

**D**

**Supplementary figure 2: A-** 40377- blood sample; **B**- 40377- liquid culture sample; **C-** 20790- blood sample; **D**- 20790 liquid culture sample.
